# Health Utility Adjusted Survival: a Composite Endpoint for Clinical Trial Designs

**DOI:** 10.1101/2024.04.08.24305511

**Authors:** Yangqing Deng, John R. de Almeida, Wei Xu

## Abstract

Many randomized trials have used overall survival as the primary endpoint for establishing non-inferiority of one treatment compared to another. However, if a treatment is non-inferior to another treatment in terms of overall survival, clinicians may be interested in further exploring which treatment results in better health utility scores for patients. Examining health utility in a secondary analysis is feasible, however, since health utility is not the primary endpoint, it is usually not considered in the sample size calculation, hence the power to detect a difference of health utility is not guaranteed. Furthermore, often the premise of non-inferiority trials is to test the assumption that an intervention provides superior quality of life or toxicity profile without compromising the survival when compared to the existing standard. Based on this consideration, it may be beneficial to consider both survival and utility when designing a trial. There have been methods that can combine survival and quality of life into a single measure, but they either have strong restrictions or lack theoretical frameworks. In this manuscript, we propose a method called HUS (Health Utility adjusted Survival), which can combine survival outcome and longitudinal utility measures for treatment comparison. We propose an innovative statistical framework as well as procedures to conduct power analysis and sample size calculation. By comprehensive simulation studies involving summary statistics from the PET-NECK trial,^1^ we demonstrate that our new approach can achieve superior power performance using relatively small sample sizes, and our composite endpoint can be considered as an alternative to overall survival in future clinical trial design and analysis where both survival and health utility are of interest.

## 1. Introduction

In many clinical studies, overall survival (OS) is used as the primary endpoint to assess efficacy of treatments. Superiority trials are used to test whether a new treatment is better than a standard or control treatment, while non-inferiority trials are used to test whether the new treatment is not unacceptably worse than control. Non-inferiority trials are especially important in circumstances where the new treatment may have other benefits (e.g., lower costs, fewer side effects, improved quality of life, or is easier to implement) compared to control, and people are only interested in showing the new treatment is not worse than control in terms of OS. When non-inferiority has been established, clinicians may be interested in further examining whether the new treatment can benefit patients more in terms of health utility.^1^ Health utility is a construct, usually ranging from 0 to 1 (although theoretically can also have negative values), that quantifies the preference for a given health state experienced by a patient at a certain time point. A higher value means a healthier state, while death usually corresponds to 0. Using health utility scores at different time point during the treatment and post-treatment, statistical analysis may be performed to compare different treatment groups’ utility scores.^2–4^ However, given that the study design is usually based on the primary endpoint of OS without considering health utility, whether there will be enough power for health utility analysis is uncertain. Also, conducting tests for OS and health utility separately may not be the most efficient, because it involves multiple testing adjustment and can lose statistical power. Hence, it may be beneficial to consider using a composite endpoint that combines survival and utility, which may lead to increased statistical power and smaller required sample sizes.

Creation of a composite endpoint of survival and utility, can also aid in clinical interpretation of non-inferiority trials where non-inferiority of survival is not the only acceptable outcome. For example, a new therapeutic intervention may be purported as offering improvements in quality of life or toxicity. However, clinicians may not be willing to sacrifice disease control to provide these other benefits. In this case, testing this new intervention in phase 3 non-inferiority trial where overall survival is the primary outcome and quality of life or toxicity is a secondary outcome may establish the intervention as non-inferior from a survival perspective and then falsely identify the new intervention as a standard of care without appropriate consideration of quality of life and toxicity. On the other hand, one may consider a situation where a patients’ preference for improved quality of life (or utility) may outweigh their desire to have non-inferior survival. In this instance, demonstration of superiority of utility may not be enough if it is associated with a significant loss of survival and the two outcomes cannot be interpreted in isolation. In this instance, a combination of both survival and utility endpoints may be needed to declare a new intervention superior.

Some methods that can combine survival and utility have been proposed and used to analyze clinical trial data, and the most commonly used method is called Q-TWiST (Quality-adjusted Time Without Symptoms of disease or Toxicity).^5–12^ Though Q-TWiST has not been commonly seen as a primary endpoint for designing new studies, researchers have derived its statistical properties as well as formulas for sample size calculations.^8^ That being said, one major issue about Q-TWiST is that it divides each patient’s status into three states (toxicity, time without symptoms and toxicity, and relapse) and uses pre-selected weights for different states. In many scenarios, with utility scores measured as continuous variables at different time points throughout the trials, it may be much more desirable to analyze them in their original scales rather than forcing to have three categories, which may likely result in loss of information and decreased statistical power.

QALY (Quality-Adjusted Life Years), of which Q-TWiST can be considered as a special case, is the most intuitive way to combine survival with utility when comparing different treatments.^5, 13– 17^ It has also been used in the field of cost utility analysis, where similar methods have been proposed and compared.^18–21^ Quality-adjusted progression-free survival, a similar concept with slightly different focus, has also been used to assess the benefits of different treatments in randomized trials.^22–24^ However, such measures have rarely been considered as a primary endpoint for designing new trials, and we are not aware of any detailly developed statistical framework or comprehensive simulation studies that demonstrate the advantages and feasibility of a quality-adjusted survival endpoint compared to the traditional survival endpoint.

With these limitations and considerations, we propose an innovative composite endpoint for combining longitudinal health utility and survival, called HUS (Health Utility adjusted Survival), with a detailed statistical testing framework as well as procedures to perform power analysis and sample size calculations. By assigning weights to health utility and survival, HUS can be modified to suit different scenarios with increased power.

This new endpoint may help better interpret the findings in clinical trials. Often non-inferiority trials are plagued with uncertainty of the efficacy of a new intervention that is statistically deemed non-inferior based mainly on survival estimates but that has not been clearly shown to be more effective from a toxicity reduction or quality of life improvement perspective. In Table 1, we provide several scenarios of how the new composite endpoint of HUS may improve interpretation of clinical trial findings if this composite endpoint was used in place of standard primary endpoints. For example, one may consider three scenarios in which a new treatment is deemed non-inferior based on a primary outcome of survival in a typical non-inferiority design where different utility scores may produce drastically different trial conclusions if a composite HUS endpoint were used. If a new intervention had lower utility than the comparator, a non-inferior trial would declare the new intervention non-inferior, when in fact, a HUS endpoint would appropriately declare the new intervention inferior. In addition, as we will show in the simulations, sufficient power may be achieved with smaller sample sizes to make statistical inferences than non-inferiority trials based on a non-inferiority margin of survival. This feature may improve the efficiency of trial conduct and arriving at meaningful conclusions with smaller samples.

**Table 1.**
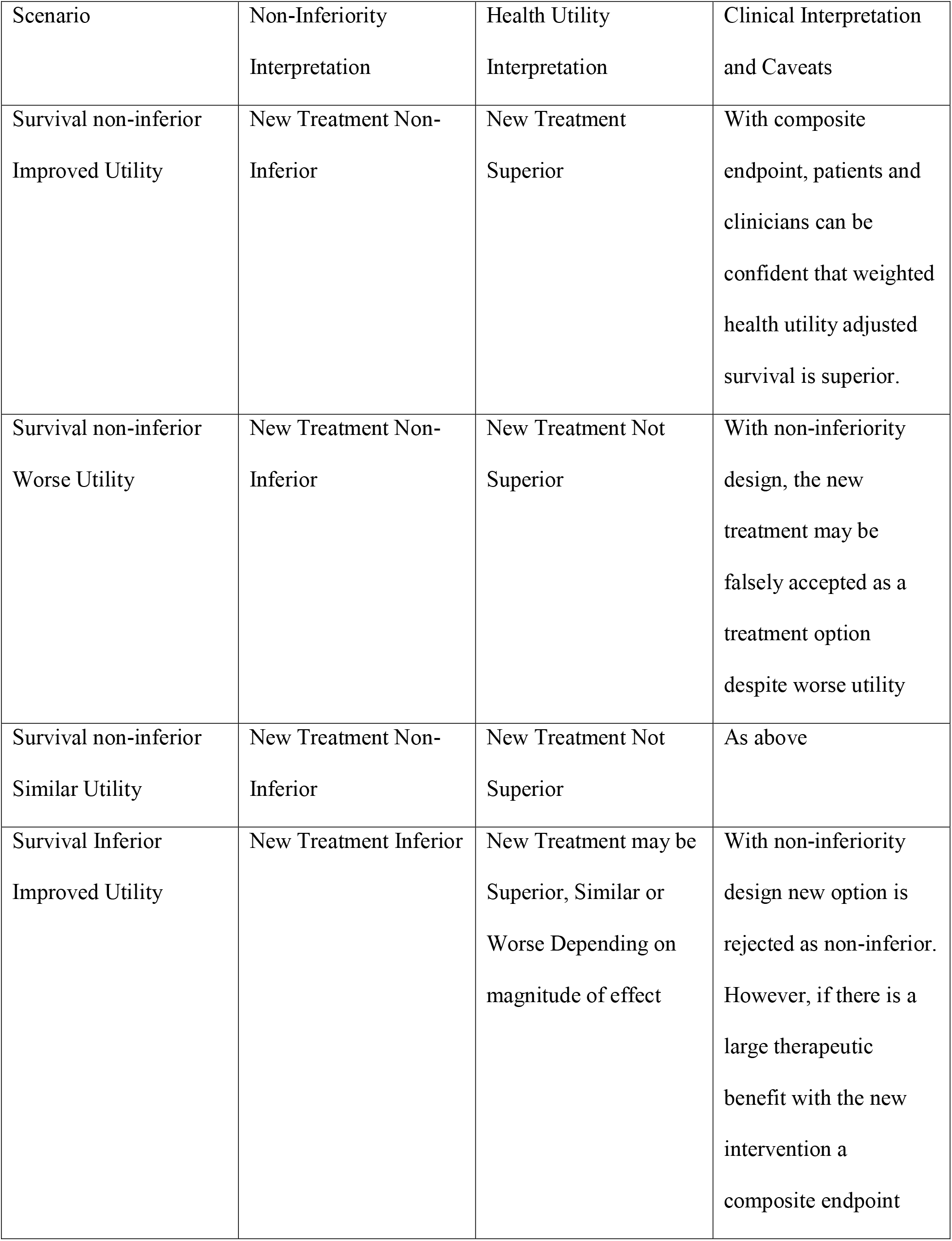
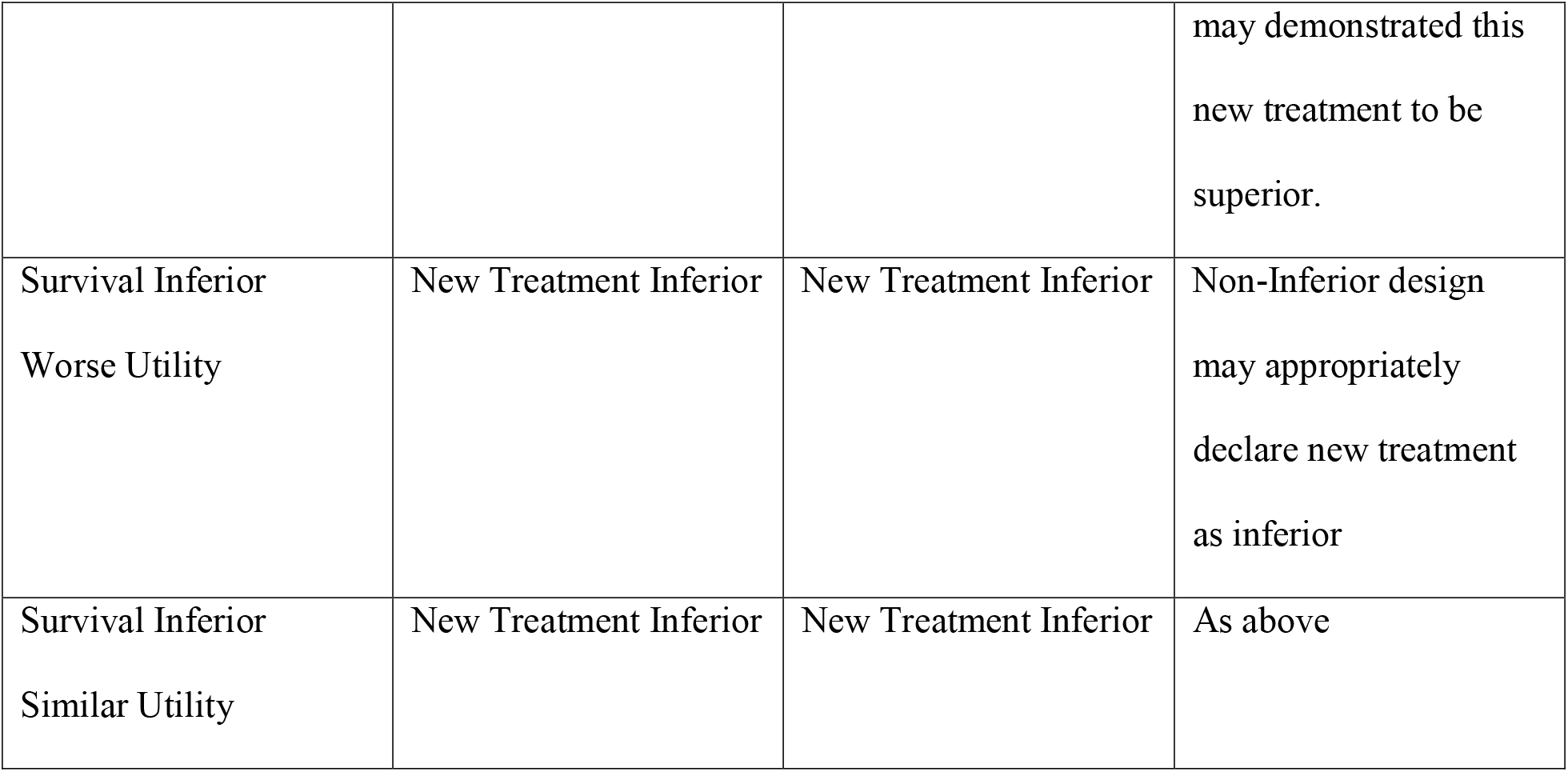
Interpretations for different scenarios of survival and utility.

This manuscript is structured as follows. In section 2, we present the methodology of the HUS endpoint, including its construction, sample size calculation and power analysis. In section 3, we use comprehensive simulation studies with various settings, including scenarios incorporating parameter estimates based on the PET-NECK trial^1^ to demonstrate the power advantage of HUS when analyzing study data and its potential to reduce required sample sizes when designing new trials. At last, we provide a discussion on the advantages, limitations and future directions for HUS in section 4.

## 2. Methods

### 2.1 Health Utility Adjusted Survival (HUS)

In this section, we describe the basic framework of Health Utility adjusted Survival (HUS). In many clinical studies, overall survival is chosen as the primary endpoint, which determines the sample size, while health utility scores are usually analyzed in the secondary analyses. To construct a composite endpoint combining survival and health utility, we can take the product of the survival curve and the utility curve, as illustrated by Figure 1.

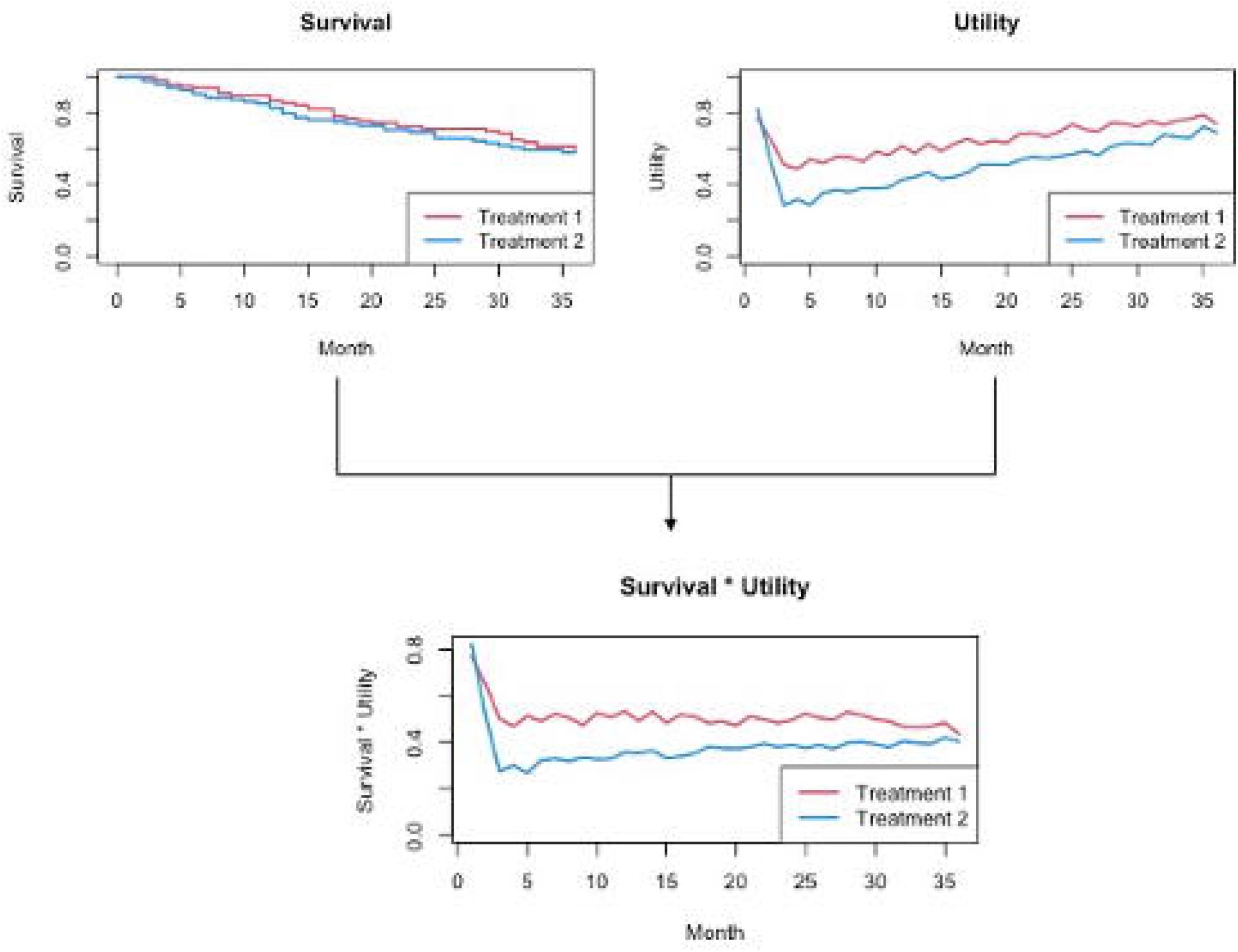

Suppose the total length of the study follow up time is *T*, and we are interested in comparing survival and health utility between treatment groups 1 and 2. We define a Q statistic to represent the HUS of each treatment group as

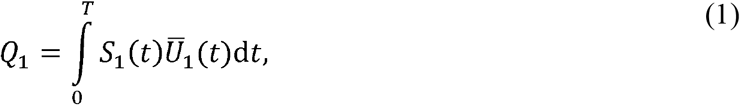

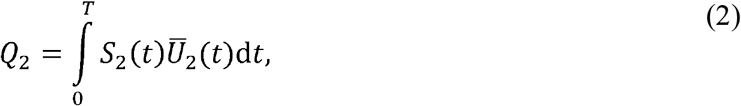

where and *S*_1_(*t*) and *Ū*_1_ (*t*) are the survival function (proportion of patients alive at *t*) and average utility score of those alive at *t* for group 1. *S*_2_(*t*) and *Ū*_2_ (*t*) are the survival function and average utility score of those alive at *t* for group 2. We propose to use the Kaplan Meier (KM) estimated survival functions *Ŝ*_1_ (*t*), *Ŝ*_2_ (*t*) to substitute *S*_1_(*t*), *S*_2_(*t*).

We can also assign weights to the survival and utility separately by defining

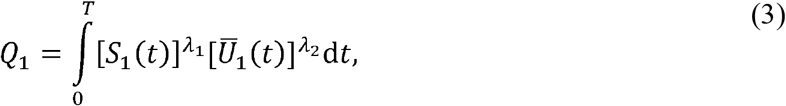

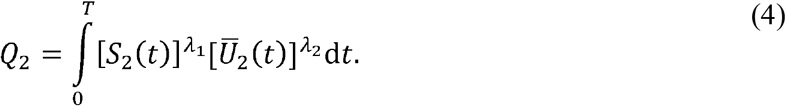

If *λ*_1_ = 0 and *λ*_2_ = 1, then *Q*_1_ and *Q*_2_ only consider the utility functions without including survival. If *λ*_1_ = 1 and *λ*_2_ = 0, then *Q*_1_ and *Q*_2_ simply calculate the areas under the survival curves without adjusting for utility. For simplicity, we suggest fixing the weight *λ*_1_ as 1, since survival is usually considered as important. *λ*_2_ can be chosen from different values (e.g., 0.5, 1, 2), and *λ*_2_ = 1 leads to the standard definition of HUS. The higher *λ*_2_ is, the more importance is assigned to health utility. For the rest of this manuscript, we focus on *λ*_1_ = 1 and *λ*_2_ = 1 unless otherwise specified. We will also show some results with various *λ*_2_ in our simulation studies and discuss its effect.

### 2.2 Hypothesis Testing

To examine the difference of HUS between two treatment groups, we can define the test statistic as

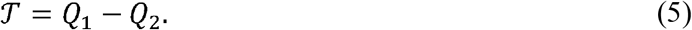

To perform a one-sided test on whether group 1 has better HUS than group 2, we can either use the bootstrap method to obtain the confidence interval of *𝒯*, or use the permutation method to obtain the distribution of *𝒯* under the null hypothesis.^25^ We can reject or accept the null hypothesis (H0: *𝒯* ≤ 0) based on bootstrap confidence intervals. Suppose groups 1 and 2 have *n*_1_ and *n*_2_ subjects respectively, and the chosen significance threshold is *α*. The bootstrap procedure can be described as follows:

1. For iteration *b*(*b*=1,…, *B*), take a bootstrap dataset from the original samples, meaning that we randomly sample *n*_1_subjects with replacement from treatment group 1 to be group 1 in the new sample, *n*_2_ subjects with replacement from treatment group 2 to be group 2 in the new sample.
2. Calculate the *𝒯* test statistic for the new sample, denoted by *𝒯*^(*b*)^.
3. After obtaining *𝒯*^(*b*)^’s (*b*=1,…,*B*), calculate the (1−*α*) confidence interval based on these *B* bootstrap samples. If the confidence interval does not contain 0, reject the null hypothesis. Note that the confidence interval should be constructed based on the test of interest (one-sided or two-sided).

The permutation procedure can be described as follows:

1. For iteration *b*(*b*=1,…, *B*), permute on the original samples to get a new permutation dataset, meaning that we randomly reassign all of the subjects into two groups with sample sizes *n*_1_ and *n*_2_.
2. Calculate the *𝒯* test statistic for the new sample, denoted by *𝒯*^(*b*)^.
3. After obtaining *𝒯*^(*b*)^’s (*b* =1,…, *B*), calculate the (1− *α*) confidence interval based on these *B* permutation samples. If the observed test statistic *𝒯* is outside the confidence interval, reject the null hypothesis.

Note that the distribution generated by bootstrap is under the alternative hypothesis, whereas the distribution generated by permutation is under the null hypothesis, which is why the former is compared with 0, while the latter is compared with the observed test statistic. Based on our experience, both bootstrap and permutation methods can control type I errors, but bootstrap tends to have slightly higher power than permutation. Hence, we focus on the bootstrap method by default. Some simulation results comparing bootstrap and permutation can be found in the supplementary materials (Table S4, Figure S1). Besides, as hinted by Glasziou et al.,^13^ Jackknife resampling can also be used to obtain the distribution of *𝒯* under the alternative hypothesis.^26, 27^ However, our past experience shows that there is little difference in terms of type I error and power when comparing the bootstrap method with Jackknife, while the distribution of *𝒯* based on bootstrap samples tends to be closer to normal. As a result, we suggest using the bootstrap method as default. In terms of the number of resamples, *B*=500 is usually sufficient for controlling type I errors and obtaining decent power. Examples showing the performance of Jackknife and evaluating the choice of *B* are also provided in the supplementary materials (Tables S4-S5, Figure S1).

### 2.3 Theoretical Properties

If we assume the survival time follows a piecewise exponential distribution, we can derive a Monte Carlo approach to calculate the variance of the test statistic, which can be used for power analysis and sample size calculation.^28^ A similar idea was used by Royston and Parmar^29^ to calculate the variance for restricted mean survival time (RMST).^30–32^

We consider a simple case with three key time points: 0 (baseline), *C* (end of surgery) and *T* (end of study). Focusing on one treatment group, suppose the survival time is piecewise exponential, with piecewise constant hazards *h*_1_,*h*_2_ for time periods 0 ∼ *C, C* ∼ *T* respectively. The utility function is piecewise linear, which starts from *A*_1_ at time 0, changes to *A*_2_ at time *C*, and then goes to *A*_3_ at time *T*. Let *X* = min (*ξ,T*), where *ξ* is the survival 1 time with cumulative hazard function *H*(*t*) and survival function *S*(*t*). We can decompose *X* as *X*_1_ + *X*_2_, where

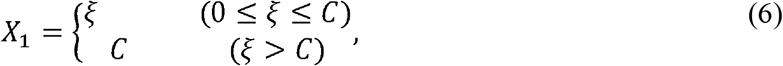

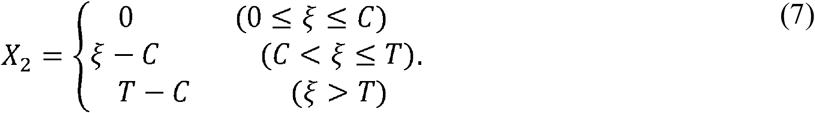

Denote 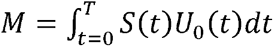 where *U*_0_ (*t*) is the base utility function for the currently considered treatment group. Write its statistic of HUS as 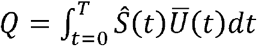. If we define

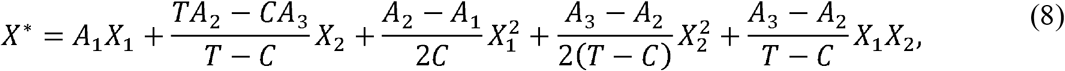

we can derive that *M* = E(*X*^*^). Following Royston & Parmar (2013), we can assume that for a specific scenario, we have

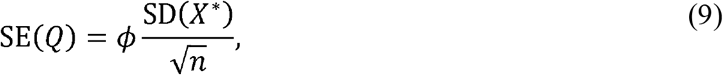

where *ϕ* is a factor no less than 1 and *n* is the sample size for the group we are currently looking at. For convenience, we call *ϕ* the variance balance factor, which takes account of the extra variance introduced into HUS by missing utility, censored survival, KM estimation, etc. SD(*X*^*^) can be calculated using the parameters, while *ϕ* can be estimated by Monte Carlo sampling. More details including the derivations are provided in the supplementary materials (Tables S2-S3). We will demonstrate in our simulations that *ϕ* is robust to different sample sizes.

Note that when two treatment groups are compared, they should have their own variance balance factors, which we denote as *ϕ*_1_ and *ϕ*_2_. Applying our assumed property to each of the groups, we have

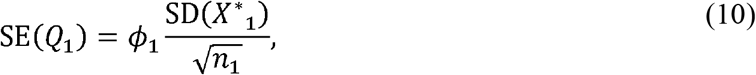

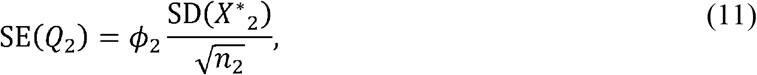

where *Q*_1_ and *Q*_2_ are the statistics of HUS for treatment groups 1 and 2, and *n*_1_,*n*_2_ are the sample sizes of the two groups. *X*^*^_1_ and *X*^*^_2_ are constructed separately for the two groups using their own parameter settings. Hence, the variance of *𝒯* = *Q*_1_ − *Q*_2_ is

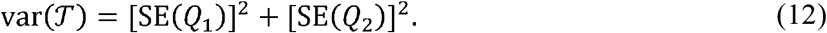

For the one-sided test, we can reject the null hypothesis if 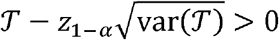.

### 2.4 Power Analysis and Sample Size Calculation

In any scenario with prespecified parameters, given different sample size, we can calculate the corresponding power of HUS using simulations. Then we can obtain a table showing different power under different sample sizes, which can be used to determine the sample size needed to achieve specific power (e.g., 80%) for a new trial. Detailed examples are provided in section 3.1.

If we assume that the special case described in section 2.3 is true, then we only need to run one simulation given a fixed sample size (e.g., 200 subjects per treatment group), which can give us estimates of *ϕ*_1_ and *ϕ*_2_. For the one-sided test where we reject the null hypothesis if 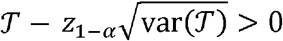, the power is

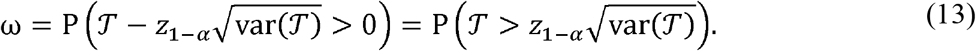

Assume *𝒯* follows *N*(*𝒯*_true_, var(*𝒯*)) and denote the power by *ω*, we have

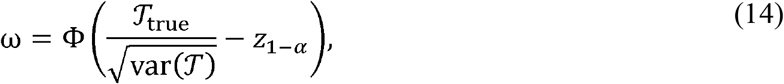

where *Φ* is the cumulative distribution function of the standard normal distribution. On the other hand, to achieve power *ω*, the required sample sizes should satisfy

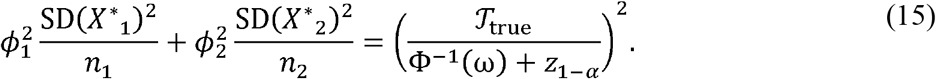

If we assume *n*_1_=*n*_2_, then the required sample size per arm is

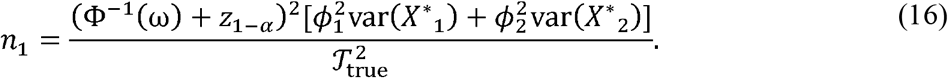

Note that it is difficult to calculate *𝒯*_true_ based on the setting of parameters. However, we can estimate it by using the average of the observed *𝒯* from our simulated samples. To summarize, in the special situation with simplified settings described in section 2.3, we can use the following procedure to calculate power yielded by a specific sample size:

1. Calculate var (*X*^*^ _1_), var (*X*^*^ _2_) based on parameter settings.
2. Simulate samples to estimate *ϕ*_1_, *ϕ*_2_ and *𝒯*_true_.
3. For each new sample size combination *n*_1_, *n*_2_, calculate SE (*Q*_1_), SE (*Q*_2_) using the estimated *ϕ*_1_, *ϕ*_2_.
4. Calculate var (*𝒯*) and power *ω*. On the other side, we can use the following procedure to calculate the sample size required to achieve specific power:

1. Calculate var (*X*^*^ _1_), var (*X*^*^ _2_) based on parameter settings.
2. Simulate samples to estimate *ϕ*_1_, *ϕ*_2_ and *𝒯*_true_.
3. Calculate *n*_1_ using the sample size formula.

### 2.5 Handling Missing Utility Scores

In clinical studies, utility scores may not be available at each time point for all subjects, while the current framework of HUS requires complete utility profiles to calculate the test statistic. The most intuitive way is to impute the utility scores. We use linear functions to fill in the utility scores using the available data. If a subject’s utility score is only available at one time point, then we use that score as the imputed utility at all other time points. This approach may seem simple, but it can be quite effective. Another method we consider is to impute the group average at each key time point (i.e., each time point at which at least one subject has their utility score recorded), and then use linear functions to fill in the other missing scores. This approach can be regarded as a combination of the cross-mean and linear interpolation methods^33^. While imputing the group average, we can also add some variation using a normal distribution with mean zero and its standard deviation equal to the standard deviation of the recorded scores at that time point. In this way, the imputed values may be closer to the true values, which may lead to increase of statistical power. It is also worth noting that many other methods are available for imputing longitudinal data, and a very recent study has compared the effects of different imputation methods and shown that most of them are similar in various scenarios, whereas trajectory mean single imputation has the best overall performance^33^. Hence, we consider trajectory mean imputation as a third method. A comparison of the three methods using simulation results is provided in the supplementary materials (Table S1), which shows that method 1 has much worse performance when the missing rate is higher, while methods 2 and 3 are not affected as much. For convenience, we use method 2 by default.

## 3. Results

### 3.1 Simulations with Simplified Settings

#### 3.1.1. Power Comparison

We conduct simulations in various scenarios to assess the performances of HUS. Suppose we are designing a randomized clinical trial with two treatment arms. The total length of study is 36 months (*T* = 36), and each patient receives surgery at 3 months (*C* = 36). The two arms are assigned to different treatment strategies to help them recover, and we are interested in comparing the two treatments in terms of both survival and health utility. Denote the true survival time, observed survival time and survival status for patient *i* from group *g* as *T*_*gi*_, *X*_*gi*_ and *δ*_*gi*_ respectively. Group 1 and group 2 have sample sizes *n*_1_ and *n*_2_. The survival data is simulated using

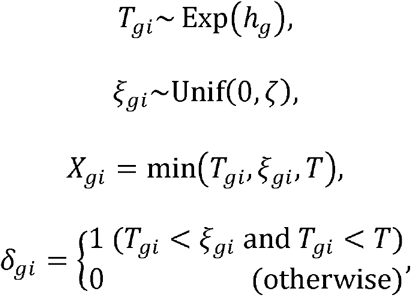

where *ζ* is chosen to control the censoring rate, denoted by *p*_censoring_. The hazard ratio of treatment 1 against treatment 2 is *h*_1_/*h*_2_.

To simulate the health utility score, we first define base utility functions for the two groups. The base utility at time *t* for group *g* can be written as

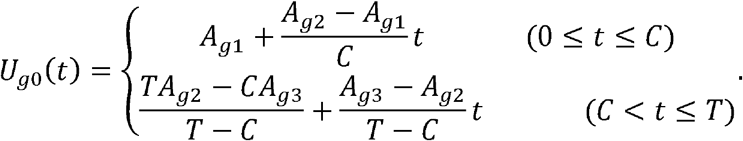

This definition means the average utility for group *g* starts from *A*_*g*1_ at baseline, changes to *A*_*g*2_ at 3 months, and then changes to *A*_*g*3_ at the end of the study. The change is piecewise linear. Our motivation for this setting is that usually a cancer patient’s health utility reaches the lowest at the end of treatment and gradually recovers after that. For patient *i* from group *g*, the health utility score at time *t*, denoted by *U*_*gi*_ (*t*), follows a normal distribution with mean *U*_*g*0_(*t*) and standard deviation 0.1.

In practice, we do not expect health utility scores to be collected at each time point. Furthermore, some of the scores scheduled to be collected may be missing. For our main simulation study, we assume that the health utility scores are only collected at *t* = 1, *C* and *T*. When *t* = 1, all subjects have their utility scores collected. When *t* = *C* or *T*, the subjects that are still being followed have their utility scores collected, while there is a *p*_missingU_ chance that the score is missing.

In this section, we focus on the situation where the two treatment groups do not have a difference in OS, which is the situation that motivated our HUS framework. Other situations (e.g., the two treatment groups differ in both OS and health utility) are explored in section 3.2 and the supplementary materials (Tables S6-S9). Table 2 shows a summary of our major scenarios. In each scenario, we compare the theoretical rejection rate using our results from section 2.4 and the empirical rejection rates of HUS using bootstrap with *B* = 500. We consider three choices of *λ*_1_ : *λ*_2_ = 1 corresponds to the standard HUS approach; *λ*_2_ = 0.5 means giving utility less weight than survival; *λ*_2_ = 2 means giving utility more weight than survival. We also examine the performance of OS-based tests. In the tables, “sup” represents the log-rank test that tests whether group 1 is superior to group 2 in terms of OS using KM estimates. “5%” and “10%” correspond to the inferiority test using the hazard ratio with margins 5% and 10% respectively. For instance, a 5% margin means we establish non-inferiority (treatment 1 is non-inferior to treatment 2 in terms of OS) if the upper bound of the 95% CI of the hazard ratio is smaller than 1.05.

**Table 2.**
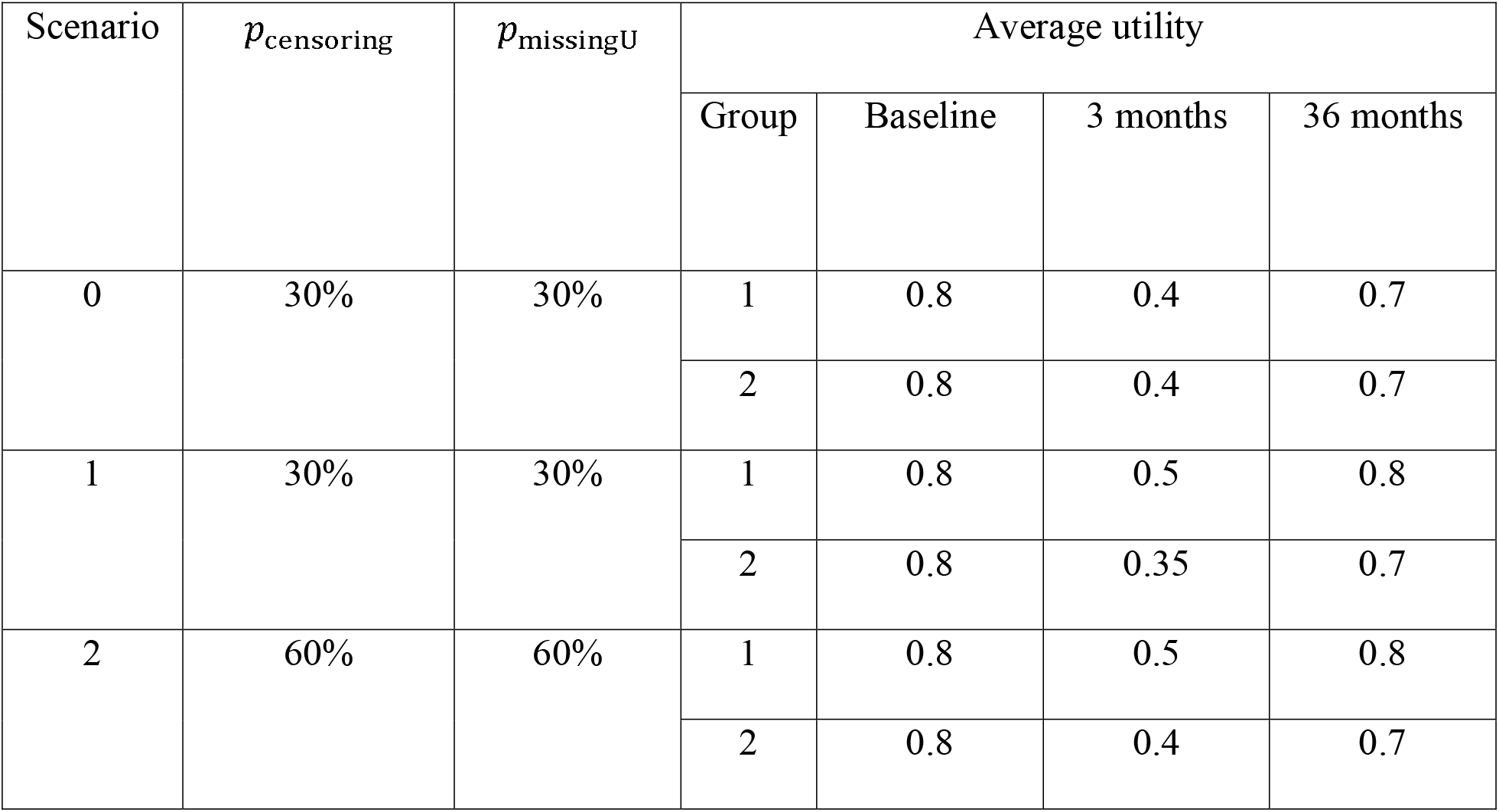
Simulation settings with different scenario.

In scenario 0, we examine the rejection rates of different methods when the two treatment groups have the same OS and health utility. As shown in Table 3, all of the superiority tests are able to control type I errors at 0.05. The rejection rates of the non-inferiority tests are power instead of type I errors, since the alternative is true (treatment 1 is not inferior to treatment 2). This is why they may be higher than 0.05.

**Table 3.** Rejection rates of different methods in scenario 0 based on 1000 replications.

| $n_1, n_2$ | HUS | | | | OS | | |
| --- | --- | --- | --- | --- | --- | --- | --- |
| | Theoretical | Bootstrap<br>($\lambda_2 = 1$) | $\lambda_2 = 0.5$ | $\lambda_2 = 2$ | Superiority<br>test | Non-inferiority<br>test with<br>margin<br>5% | Non-inferiority<br>test with<br>margin<br>10% |
| 50 | 0.052 | 0.052 | 0.050 | 0.053 | 0.056 | 0.040 | 0.050 |
| 100 | 0.052 | 0.054 | 0.053 | 0.051 | 0.068 | 0.051 | 0.073 |
| 150 | 0.053 | 0.047 | 0.052 | 0.048 | 0.054 | 0.042 | 0.079 |
| 200 | 0.053 | 0.052 | 0.046 | 0.048 | 0.053 | 0.050 | 0.092 |
| 500 | 0.054 | 0.049 | 0.051 | 0.050 | 0.045 | 0.083 | 0.184 |

In scenario 1, we compare the power of different methods when treatment group 1 has better health utility than treatment group 2. For the theoretical power analysis, firstly, we run one Simulation with *n*_1_ = *n*_2_ = 200 and 4000 replications to obtain the estimates *ϕ*_1_ = 1.07, *ϕ*_2_ = 1.12, *𝒯*_true_ = 3.11. Then we can calculate the power of different 1 sample sizes. For the other methods such as bootstrap *λ*_2_ = 1, *λ*_2_ = 0.5, and *λ*_2_ = 2, we need to simulate new datasets (200 replications) with different sample sizes to get the empirical power. Note that *ϕ*_1_, *ϕ*_2_ and *𝒯*_true_ are quite robust to different sample sizes. For example, if we use *n*_1_ = *n*_2_ = 500, the obtained estimates are *ϕ*_1_ = 1.06, *ϕ*_2_ = 1.11, *𝒯*_true_ = 3.11, which is very close to the scenario of *n*_1_ = *n*_2_ = 200. More results regarding the variance balance factors are provided in the supplementary materials (Tables S2-S3).

As shown in Table 4, the bootstrap method with *λ*_2_ = 1 performs close to the theoretical results, which makes sense since the theoretical results are based on the standard HUS with *λ*_2_ = 1. Larger *λ*_2_ tends to lead to higher power by giving more weight to utility than survival. This is also expected because the two groups only differ in terms of utility. Meanwhile, the superiority and non-inferiority tests based on OS have little power since there is no real difference in the two group’s OS. We also calculate the power corresponding to different sample sizes using our theoretical results and plot the power curves in Figure 2. For scenario 1, to achieve 80% power, using HUS as the endpoint only requires 85 subjects per arm.

**Table 4.** Power comparison of different methods in scenarios 1 and 2 based on 200 replications.

| Scenario 1 |  |  |  |  |  |  |  |
| --- | --- | --- | --- | --- | --- | --- | --- |
| $n_1, n_2$ | HUS | | | | OS | | |
| | Theoretical | Bootstrap<br>($\lambda_2 = 1$) | $\lambda_2 = 0.5$ | $\lambda_2 = 2$ | Superiority<br>test | Non-inferiority<br>test with<br>margin<br>5% | Non-inferiority<br>test with<br>margin<br>10% |
| 50 | 0.61 | 0.56 | 0.28 | 0.9 | 0.05 | 0.04 | 0.05 |
| 100 | 0.86 | 0.85 | 0.44 | 1 | 0.05 | 0.05 | 0.07 |
| 150 | 0.95 | 0.95 | 0.59 | 1 | 0.05 | 0.06 | 0.1 |
| 200 | 0.99 | 1 | 0.71 | 1 | 0.06 | 0.04 | 0.06 |
| Scenario 2 |  |  |  |  |  |  |  |
| $n_1, n_2$ | HUS | | | | OS | | |
| | Theoretical | Bootstrap<br>($\lambda_2 = 1$) | $\lambda_2 = 0.5$ | $\lambda_2 = 2$ | Superiority<br>test | Non-inferiority<br>test with<br>margin<br>5% | Non-inferiority<br>test with<br>margin<br>10% |
| 50 | 0.42 | 0.42 | 0.2 | 0.76 | 0.05 | 0.04 | 0.06 |
| 100 | 0.65 | 0.67 | 0.31 | 0.94 | 0.06 | 0.06 | 0.08 |
| 150 | 0.80 | 0.82 | 0.42 | 0.97 | 0.06 | 0.05 | 0.1 |
| 200 | 0.89 | 0.92 | 0.48 | 1 | 0.05 | 0.05 | 0.06 |

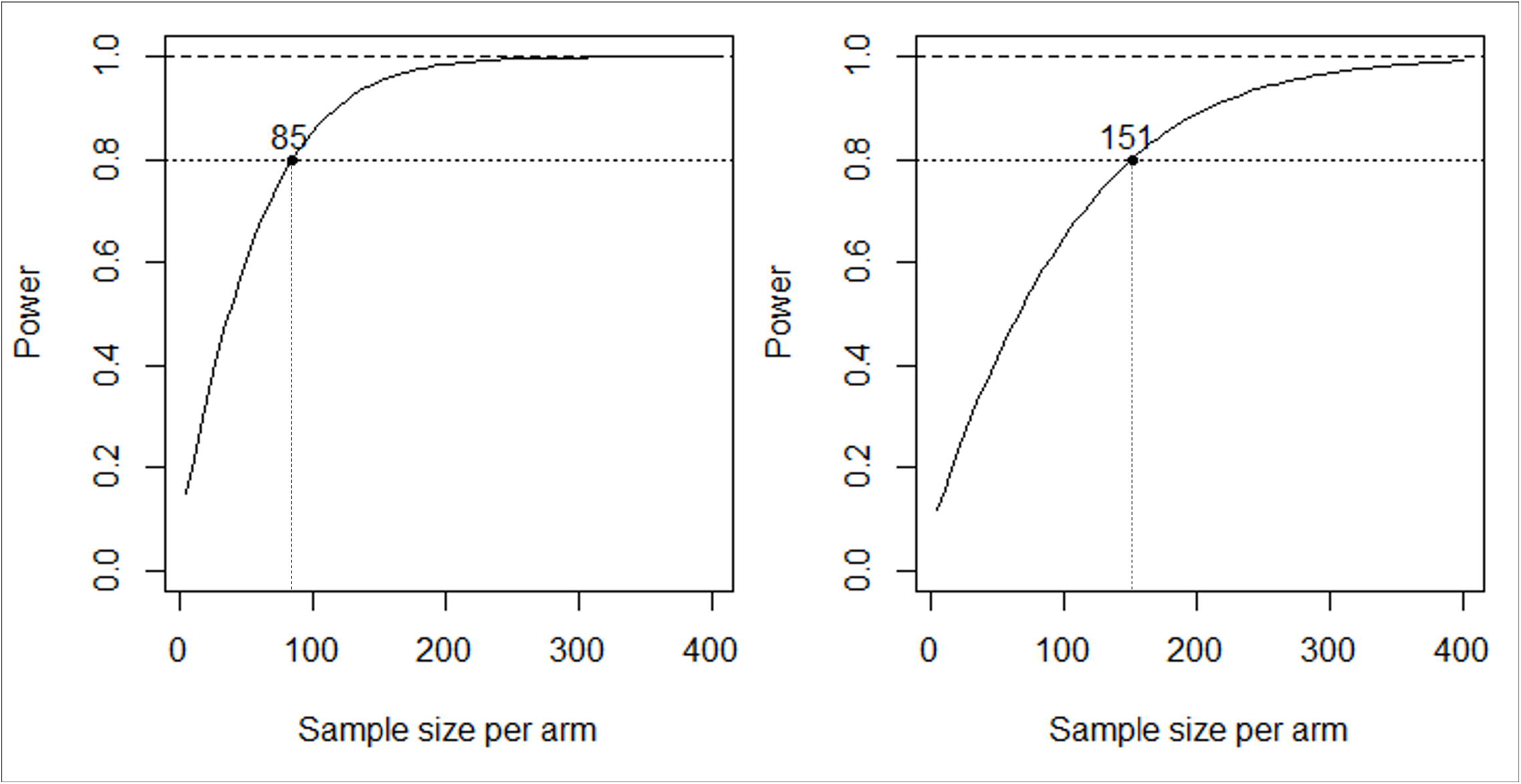

In scenario 2, we increase the censoring rate to 60% and missing rates to 60%, and reduce the difference between the two group’s health utility scores. As shown in Table 4 and Figure 2, results are very similar to those in scenario 1. Again, HUS is able to obtain decent power with relatively small sample sizes while the superiority and non-inferiority tests struggle to find enough evidence to show treatment 1’s benefit compared to treatment 2. If we design a trial based on HUS with the assumptions in scenario 2, we only need to have 151 patients in each treatment group.

We would like to point out that even though choosing a larger *λ*_2_ may seem to have higher power in the above scenarios, it may not always be a good choice, especially when there is a difference in OS. We recommend using *λ*_2_ = 1 as default, though it can be modified depending on the knowledge of the two treatments (e.g., whether treatment 1 is likely to have better OS than treatment 2).

#### 3.1.2 Sample Size Calculation

In this subsection, we use our developed sample size calculation formulas to calculate sample sizes needed for the composite endpoint, and the standard formulas to calculate sample sizes needed for basic survival endpoint (implemented in PASS 2023, v23.0.2 with the one-sided log-rank test), to further demonstrate the advantage of HUS. Following scenario 1 from 3.1.1, where treatment 1 has better utility than treatment 2, we consider four different cases. In the first case, there is no survival difference, which is consistent with the focus of this manuscript, and the endpoint overall survival does not have power. In the second case, we assume that treatment 1 has better survival than treatment 2, while in the third case, we assume that treatment 2 has better survival. In the last case, we assume treatment 1 has better survival, but there is no difference in utility, and the utility function is the same as that in scenario 0. As shown in Table 5, with scenario 1’s utility functions, when *h*_1_ is smaller than *h*_2_, meaning that treatment 1 has better OS than treatment 2, the required sample size for HUS is decreased, which makes sense because the difference in HUS is larger. When *h*_1_ is larger than *h*_2_, meaning that treatment 1 has worse OS than treatment 2, the required sample size for HUS is increased. Nevertheless, the numbers are still much smaller than those calculated for overall survival. If there is no utility difference, HUS will require more subjects than overall survival, which is expected, though the difference is not as big. These results show again that using the composite endpoint may help greatly reduce the required sample size to detect a significant difference when two treatments differ in utility.

**Table 5.** Sample size calculation under a significance level of 0.05. When there is a utility difference, utility functions from scenario 1 are used. When there is no utility difference, utility functions from scenario 0 are used.

| Utility: 1>2; survival: 1=2 ($h_1/h_2 = 1$) | | |
| --- | --- | --- |
| Targeted power | Sample size requirement for each arm |  |
|  | HUS | OS |
| 70% | 65 | / |
| 80% | 85 | / |
| 90% | 118 | / |
| Utility: 1>2; survival: 1>2 ($h_1/h_2 = 0.9$) | | |
| Targeted power | Sample size requirement for each arm |  |
|  | HUS | OS |
| 70% | 47 | 1710 |
| 80% | 62 | 2247 |
| 90% | 85 | 3112 |
| Utility: 1>2; survival: 1<2 ($h_1/h_2 = 1.1$) | | |
| Targeted power | Sample size requirement for each arm |  |
|  | HUS | OS |
| 70% | 78 | 2243 |
| 80% | 103 | 2946 |
| 90% | 143 | 4080 |

| Utility: 1=2; survival: 1>2 ($h_1/h_2 = 0.7$) | | |
| --- | --- | --- |
| Targeted power | Sample size requirement for each arm |  |
|  | HUS | OS |
| 70% | 184 | 138 |
| 80% | 242 | 180 |
| 90% | 335 | 249 |

### 3.3. Simulations with Real Data Estimates

To demonstrate the benefit of HUS in a more practical scenario, we conduct additional simulations with average utility scores and the hazard ratio mimicking the summary data provided in a real randomized trial PET-NECK.^1^ PET-NECK is a randomized phase III non-inferiority trial that compares Positron emission tomography-computerized tomography-guided watch-and-wait policy (PET-CT) with planned neck dissection (planned ND) for head and neck cancer patients. The two-year overall survival rates of the two treatment groups (PET-CT and planned ND) with 282 subjects per arm, are 84.9% and 81.5% respectively, which leads to a hazard ratio of 0.80. We conducted simulations utilizing the parameter setting to emulate the survival times in PET-NECK. Figure 3 shows the average utility scores at different time points in the study, with the maximum time being 24 months. Hence, in this scenario, we set *T* = 24 and define the base utility functions following the observed average utility scores. We also record the utilities at baseline and months 1, 3, 6, 12, 24 with 30% missing rate. Note that this scenario does not fall into the framework of section 2.3, and thus we cannot apply our theoretical results directly to calculate the power and sample sizes. However, obtaining the empirical results is similar to what we describe in section 3.1.

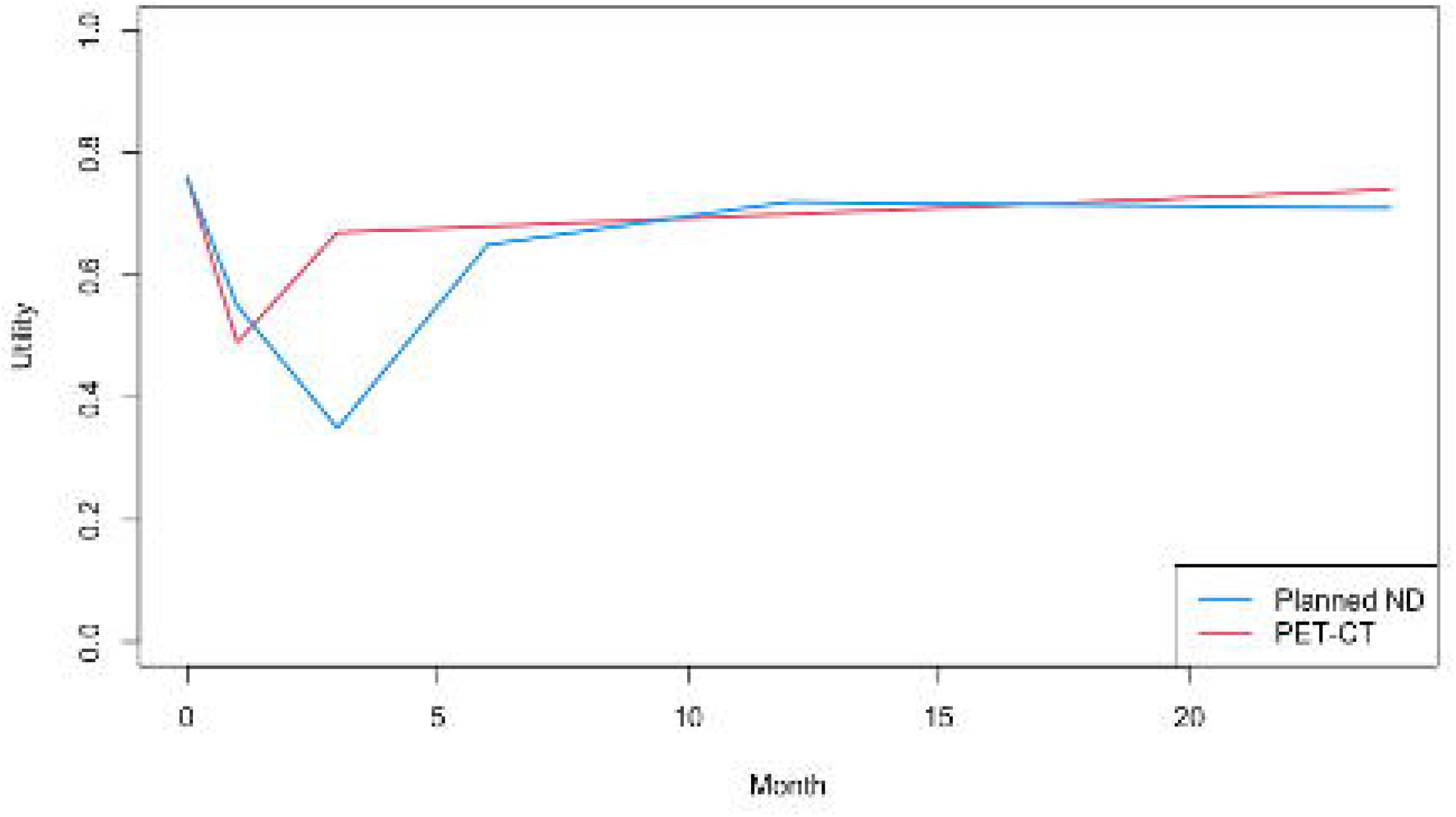

As shown in Table 6, with the two groups differing in both OS and health utility, the superiority test based on HUS still has much higher power than the superiority and non-inferiority tests based on OS. We would only need 200 subjects per arm to achieve 80% power of showing PET-CT has better HUS than planned ND, which is fewer than the subjects in the original study which were based on OS comparison. In terms of weighting, *λ*_2_ = 2 again leads to higher power, while *λ*_2_ = 0.5 has lower power compared to the standard HUS. Nevertheless, in certain scenarios, especially if the difference in health utility is small, using a larger *λ*_2_ may not be as beneficial. More results, including scenarios where there is no difference in health utility, are available in the supplementary materials (Tables S6-S9, Figures S2-S4).

**Table 6.** Power comparison for simulations using real data estimates.

|  |  |  |
| --- | --- | --- |
| $n_1, n_2$ | HUS | OS |

| | Bootstrap<br>($\lambda_2 = 1$) | $\lambda_2 = 0.5$ | $\lambda_2 = 2$ | Superiority<br>test | Non-inferiority<br>test with<br>margin 5% | Non-inferiority<br>test with<br>margin 10% |
| --- | --- | --- | --- | --- | --- | --- |
| 50 | 0.34 | 0.26 | 0.47 | 0.09 | 0.1 | 0.12 |
| 100 | 0.54 | 0.4 | 0.69 | 0.14 | 0.17 | 0.22 |
| 150 | 0.74 | 0.54 | 0.84 | 0.19 | 0.24 | 0.34 |
| 200 | 0.82 | 0.61 | 0.97 | 0.2 | 0.25 | 0.33 |
| 282 | 0.94 | 0.76 | 0.99 | 0.27 | 0.36 | 0.44 |

## 4. Discussion

We have presented a methodological framework to compare two treatment groups using HUS as a composite endpoint combining survival and health utility. As demonstrated by our comprehensive simulation studies, when there is a difference in health utility, HUS has a significant power advantage over the statistical tests based on OS endpoint, meaning that using HUS as an endpoint for new trials may require much smaller sample sizes to achieve decent power. We have also demonstrated two different procedures (theoretical and empirical approaches) to conduct power analysis and sample size calculation with specified parameters. When the model assumptions are met, the two procedures yield similar results.

There are several different options when applying HUS. We recommend using bootstrap given its popularity as well as its convenience of constructing confidence intervals for the test statistic, though permutation may be theoretically more appropriate for testing the null hypothesis, since it can obtain the null distribution of the test statistic. In terms of weighting on survival and utility, we recommend choosing weights *λ*_1_ = *λ*_2_ = 1 as default. Using a larger *λ*_2_ may increase the power in certain scenarios, especially when the two treatment groups differ in health utility but not in OS. However, it may not be beneficial when there is no difference in health utility. A possible way to combine different weighting options without having to choose one is to apply an idea similar to the aSPU test.^34, 35^

Note that the theoretical properties we have presented are based on assumptions by analogy with the assumptions used by Royston and Parmar,^29^ though we have shown the validity of our theoretical results in our simulations. In the future, we may explore the asymptotic properties with relaxed assumptions (e.g., the survival times do not have to be piece-wise exponential), which would be helpful when designing or analyzing trials where our previously used assumptions are likely to be violated. We would also like to point out that the linear imputation method we use to fill in the utility scores may be problematic in some cases, especially if the scores are only recorded at a few time points and the missing rate is high. We may consider other imputation methods or modifying the definition of HUS so that it does not require complete utility score profiles as input.^36, 37^ Besides that, given the possible drawbacks brought by KM estimates, sometimes it may be beneficial to apply other models, including the flexible parametric model for survival analysis.^38^

Another possible direction worth exploring is to take different functions of the utility score into consideration. One special case is that in many clinical studies, multiple measures of health status are recorded. There are various ways to combine different measures into a single utility score.^39–41^ Extending HUS to be able to handle any function of utility may potentially increase power and help us gain insight on how the utility is different in different treatment groups. Furthermore, considering utility may have different importance at different time points, we may assign different weights across time. For example, having a better utility score at the later stage of the study, which means the patients have recovered better, may be more important than having a better utility score at the end of surgery. In such case, we can consider giving higher weights to later time points, and the resulted HUS may provide a clearer picture of which treatment is more beneficial for recovery.

## Data Availability

All data produced in the present study are available upon reasonable request to the authors

## 5. Data Availability

R code for our simulation studies and summary data of health utility are available at https://github.com/yangq001/HUS.

## Acknowledgments

The authors would like to acknowledge the contributions of Dr. Hisham Mehanna (Institute of Head and Neck Studies and Education, University of Birmingham) and Dr. Sue Yom (Department of Radiation Oncology, University of California) for clinical insights and discussion.

