## Supplementary Materials for "Health Utility Adjusted Survival: a Composite Endpoint for Clinical Trial Designs"

### Appendix A, Proof of Theoretical Properties

For this proof, we focus on one treatment group. Consider three key time points: 0 (baseline), $C$ (end of surgery) and $T$ (end of study). Assume the survival time is piecewise exponential, with piecewise constant hazards $h_{1}, h_{2}$ for time periods $0\sim C$, $C\sim T$ respectively. As a result, the cumulative hazard function is

$$H\left( t \right)=\left\{ \begin{aligned} h_{1}t (0\leq t\leq C) \\ h_{1}C+h_{2}(t-C) (C<t\leq T) \end{aligned} \right..$$

Since $H\left( t \right)=-ln[S\left( t \right)]$ and $S\left( t \right)=exp[-H\left( t \right)]$, we have

$$S\left( t \right)=\left\{ \begin{aligned} e^{-h_{1}t} (0\leq t\leq C) \\ e^{-h_{1}C-h_{2}(t-C)} (C<t\leq T) \end{aligned} \right..$$

Assume the base utility function $U_{0}\left( t \right)$ is piecewise linear, with

$$U_{0}\left( t \right)=\left\{ \begin{aligned} \frac{\left( t-0 \right)*A_{2}+\left( C-t \right)*A_{1}}{C} (0\leq t\leq C) \\ \frac{\left( t-C \right)*A_{3}+\left( T-t \right)*A_{2}}{T-C} (C<t\leq T) \end{aligned} \right.,$$

which is equivalent to

$$U_{0}\left( t \right)=\left\{ \begin{aligned} A_{1}+\frac{A_{2}-A_{1}}{C}t (0\leq t\leq C) \\ \frac{TA_{2}-CA_{3}}{T-C}+\frac{A_{3}-A_{2}}{T-C}t (C<t\leq T) \end{aligned} \right..$$

With definitions $M=\int_{t=0}^{T} S\left( t \right)U_{0}\left( t \right)dt$ and $Q=\int_{t=0}^{T} \hat{S}\left( t \right)U\left( t \right)dt$, following Royston and Parmar^1^, if we can write $M=E(X^{*})$ for some variable $X^{*}$, we can assume

$$\mathrm{SE}\left( Q \right)=\phi\frac{\mathrm{SD}\left( X^{*} \right)}{\sqrt{n}},$$

with $\phi$ as a factor no less than 1. If we can calculate the variance of $X^{*}$ based on model parameters and estimate $\phi$ using one simulation (with a large sample size), we can calculate $\mathrm{SE}\left( Q \right)=\phi\mathrm{SD}\left( X^{*} \right)/\sqrt{n}$ for any given sample size $n$, without having to conduct additional simulate.

**Constructing** $\boldsymbol{X}^{\boldsymbol{*}}$

Let $X=min(\xi,T)$, where $\xi$ is the survival time with cumulative hazard function $H\left( t \right)$ and survival function $S\left( t \right)$. We can calculate

$$W_{1}=\int_{t=0}^{C} S\left( t \right)dt=\int_{t=0}^{C} e^{-h_{1}t}dt=\frac{1-e^{-h_{1}C}}{h_{1}},$$

$$W_{2}=\int_{t=C}^{T} S\left( t \right)dt=\int_{t=C}^{T} e^{-h_{1}C-h_{2}(t-C)}dt=\frac{e^{-h_{1}C}-e^{-h_{1}C-h_{2}(T-C)}}{h_{2}}.$$

We also have

$$L_{1}=\int_{t=0}^{C} tS\left( t \right)dt=\int_{t=0}^{C} te^{-h_{1}t}dt=-\frac{1}{h_{1}}\int_{t=0}^{C} tde^{-h_{1}t}=-\frac{1}{h_{1}}\left\{ Ce^{-h_{1}C}-\int_{t=0}^{C} e^{-h_{1}t}dt \right\}=-\frac{Ce^{-h_{1}C}-W_{1}}{h_{1}},$$

and

$$L_{2}=\int_{t=C}^{T} tS\left( t \right)dt=e^{-h_{1}C+h_{2}C}\int_{t=C}^{T} te^{-h_{2}t}dt=-\frac{e^{-h_{1}C+h_{2}C}}{h_{2}}\int_{t=C}^{T} tde^{-h_{2}t}=-\frac{e^{-h_{1}C+h_{2}C}}{h_{2}}\left\{ Te^{-h_{2}T}-Ce^{-h_{2}C}-\int_{t=C}^{T} e^{-h_{2}t}dt \right\}=-\frac{Te^{-h_{1}C+h_{2}C-h_{2}T}-Ce^{-h_{1}C}-W_{2}}{h_{2}}.$$

Hence,

$$EX=\int_{t=0}^{T} S\left( t \right)dt=W_{1}+W_{2},$$

$$EX^{2}=2\int_{t=0}^{T} tS\left( t \right)dt=2\left( L_{1}+L_{2} \right)=2\left( \frac{W_{1}-Ce^{-h_{1}C}}{h_{1}}+\frac{W_{2}-Te^{-h_{1}C+h_{2}C-h_{2}T}+Ce^{-h_{1}C}}{h_{2}} \right),$$

$$\mathrm{Var}\left( X \right)=2\left( L_{1}+L_{2} \right)-\left( W_{1}+W_{2} \right)^{2}.$$

Define

$$M=\int_{t=0}^{T} S\left( t \right)U_{0}\left( t \right)dt.$$

We know

$$S\left( t \right)U_{0}\left( t \right)=\left\{ \begin{aligned} A_{1}S\left( t \right)+\frac{A_{2}-A_{1}}{C}tS\left( t \right) (0\leq t\leq C) \\ \frac{TA_{2}-CA_{3}}{T-C}S\left( t \right)+\frac{A_{3}-A_{2}}{T-C}tS\left( t \right) (C<t\leq T) \end{aligned} \right..$$

Hence,

$$\int_{t=0}^{C} S\left( t \right)U_{0}\left( t \right)dt=A_{1}W_{1}+\frac{A_{2}-A_{1}}{C}L_{1},$$

$$\int_{t=C}^{T} S\left( t \right)U_{0}\left( t \right)dt=\frac{TA_{2}-CA_{3}}{T-C}W_{2}+\frac{A_{3}-A_{2}}{T-C}L_{2},$$

$$M=A_{1}W_{1}+\frac{TA_{2}-CA_{3}}{T-C}W_{2}+\frac{A_{2}-A_{1}}{C}L_{1}+\frac{A_{3}-A_{2}}{T-C}L_{2}.$$

Define

$$X_{1}=\left\{ \begin{aligned} \xi(0\leq\xi\leq C) \\ C (\xi>C) \end{aligned} \right.,$$

$$X_{2}=\left\{ \begin{aligned} 0 (0\leq\xi\leq C) \\ \xi-C (C<\xi\leq T) \\ T-C (\xi>T) \end{aligned} \right..$$

Then $X=X_{1}+X_{2}$.

$$EX_{1}=\int_{t=0}^{C} S\left( t \right)dt=W_{1},$$

$$EX_{2}=EX-EX_{1}=W_{2}.$$

Also,

$$EX_{1}^{2}=2L_{1}.$$

Since $X_{1}X_{2}=CX_{2}$ for any $\xi$ (if $0\leq\xi\leq C$, then $X_{1}X_{2}=\xi\cdot0=C\cdot0$),

$$EX_{1}X_{2}=CEX_{2}=CW_{2}.$$

Since $E{(X_{1}+X_{2})}^{2}=EX_{1}^{2}+2EX_{1}X_{2}+EX_{2}^{2}=2\left( L_{1}+L_{2} \right),$ we have

$$EX_{2}^{2}=2\left( L_{1}+L_{2} \right)-2L_{1}-2CW_{2}=2L_{2}-2CW_{2},$$

$$E\left( X_{1}^{2}+X_{2}^{2} \right)=2\left( L_{1}+L_{2} \right)-2CW_{2}.$$

Now we can write

$$M=A_{1}EX_{1}+\frac{TA_{2}-CA_{3}}{T-C}EX_{2}+\frac{A_{2}-A_{1}}{2C}EX_{1}^{2}+\frac{A_{3}-A_{2}}{2\left( T-C \right)}\left( EX_{2}^{2}+2EX_{1}X_{2} \right)$$

$$=E\left\{ A_{1}X_{1}+\frac{TA_{2}-CA_{3}}{T-C}X_{2}+\frac{A_{2}-A_{1}}{2C}X_{1}^{2}+\frac{A_{3}-A_{2}}{2\left( T-C \right)}X_{2}^{2}+\frac{A_{3}-A_{2}}{T-C}X_{1}X_{2} \right\}$$

Hence, if we define

$$X^{*}=A_{1}X_{1}+\frac{TA_{2}-CA_{3}}{T-C}X_{2}+\frac{A_{2}-A_{1}}{2C}X_{1}^{2}+\frac{A_{3}-A_{2}}{2\left( T-C \right)}X_{2}^{2}+\frac{A_{3}-A_{2}}{T-C}X_{1}X_{2},$$

we have $M=E(X^{*})$.

**Finding the variance of** $\boldsymbol{X}^{\boldsymbol{*}}$

To calculate $\mathrm{Var}\left( X^{*} \right)$, we need to derive higher moments of $X_{1}$ and $X_{2}$. Similar to $EX_{1}^{2}=2L_{1}$, assuming $K_{1}=\int_{t=0}^{C} t^{2}S\left( t \right)dt$, $K_{2}=\int_{t=C}^{T} t^{2}S\left( t \right)dt$, $O_{1}=\int_{t=0}^{C} t^{3}S\left( t \right)dt$, $O_{2}=\int_{t=C}^{T} t^{3}S\left( t \right)dt$, we can derive

$$K_{1}=\frac{2L_{1}-C^{2}e^{-h_{1}C}}{h_{1}},$$

$$O_{1}=\frac{3K_{1}-C^{3}e^{-h_{1}C}}{h_{1}},$$

$$K_{2}=\frac{2L_{2}+C^{2}e^{-h_{1}C}-T^{2}e^{-h_{1}C+h_{2}C-h_{2}T}}{h_{2}},$$

$$O_{2}=\frac{3K_{1}-C^{3}e^{-h_{1}C}}{h_{1}},$$

since

$$K_{2}=\int_{t=C}^{T} t^{2}S\left( t \right)dt=e^{-h_{1}C+h_{2}C}\int_{t=C}^{T} t^{2}e^{-h_{2}t}dt=-\frac{e^{-h_{1}C+h_{2}C}}{h_{2}}\int_{t=C}^{T} t^{2}de^{-h_{2}t}=-\frac{e^{-h_{1}C+h_{2}C}}{h_{2}}\left\{ T^{2}e^{-h_{2}T}-C^{2}e^{-h_{2}C}-2\int_{t=C}^{T} te^{-h_{2}t}dt \right\}=\frac{2L_{2}+C^{2}e^{-h_{1}C}-T^{2}e^{-h_{1}C+h_{2}C-h_{2}T}}{h_{2}},$$

and

$$O_{2}=\int_{t=C}^{T} t^{3}S\left( t \right)dt=e^{-h_{1}C+h_{2}C}\int_{t=C}^{T} t^{3}e^{-h_{2}t}dt=-\frac{e^{-h_{1}C+h_{2}C}}{h_{2}}\int_{t=C}^{T} t^{3}de^{-h_{2}t}=-\frac{e^{-h_{1}C+h_{2}C}}{h_{2}}\left\{ T^{3}e^{-h_{2}T}-C^{3}e^{-h_{2}C}-3\int_{t=C}^{T} t^{2}e^{-h_{2}t}dt \right\}=\frac{3K_{2}+C^{3}e^{-h_{1}C}-T^{3}e^{-h_{1}C+h_{2}C-h_{2}T}}{h_{2}}.$$

Similarly, we can also derive

$$E{X_{1}}^{3}=3K_{1},$$

$$E{X_{1}}^{4}=4K_{2}.$$

Note that ${X_{1}}^{j}{X_{2}}^{k}=C^{j}{X_{2}}^{k}$ for $k>0$. Hence, we have

$$E{X_{1}}^{j}{X_{2}}^{k}=C^{j}E{X_{2}}^{k}.$$

We also know that $E\left( X_{1}+X_{2} \right)^{3}=3(K_{1}+K_{2})$, $E\left( X_{1}+X_{2} \right)^{4}=4\left( O_{1}+O_{2} \right),$ so

$$E{X_{2}}^{3}=E\left( X_{1}+X_{2} \right)^{3}-E{X_{1}}^{3}-3E{X_{1}}^{2}X_{2}-3EX_{1}{X_{2}}^{2}=3\left( K_{1}+K_{2} \right)-3K_{1}-3C^{2}W_{2}-6C\left( L_{2}-CW_{2} \right)$$

$$=3K_{2}-6CL_{2}+3C^{2}W_{2},$$

$$E{X_{2}}^{4}=E\left( X_{1}+X_{2} \right)^{4}-E{X_{1}}^{4}-4E{X_{1}}^{3}X_{2}-6E{X_{1}}^{2}{X_{2}}^{2}-4EX_{1}{X_{2}}^{3}=4\left( O_{1}+O_{2} \right)-4O_{1}-4C^{3}W_{2}-12C^{2}\left( L_{2}-CW_{2} \right)-4C\left[ 3\left( K_{1}+K_{2} \right)-3K_{1}-6CL_{2}+3C^{2}W_{2} \right]=4O_{2}-4C^{3}W_{2}+12C^{2}L_{2}-12CK_{2}.$$

To summarize, we have

$$EX_{1}=W_{1},$$

$$EX_{2}=W_{2},$$

$$EX_{1}^{2}=2L_{1},$$

$$EX_{2}^{2}=2L_{2}-2CW_{2},$$

$$E{X_{1}}^{3}=3K_{1},$$

$$E{X_{2}}^{3}=3K_{2}-6CL_{2}+3C^{2}W_{2}$$

$$E{X_{1}}^{4}=4K_{2},$$

$$E{X_{2}}^{4}=4O_{2}-4C^{3}W_{2}+12C^{2}L_{2}-12CK_{2},$$

$$E{X_{1}}^{j}{X_{2}}^{k}=C^{j}E{X_{2}}^{k} \left( k>0 \right).$$

As a result, denoting $X^{*}=k_{1}X_{1}+k_{2}X_{2}+k_{3}X_{1}^{2}+k_{4}X_{2}^{2}+k_{5}X_{1}X_{2}$, we have

$$EX^{*}=k_{1}EX_{1}+k_{2}EX_{2}+k_{3}EX_{1}^{2}+k_{4}EX_{2}^{2}+k_{5}EX_{1}X_{2},$$

$$E{X^{*}}^{2}={k_{1}}^{2}EX_{1}^{2}+{k_{2}}^{2}EX_{2}^{2}+{k_{3}}^{2}E{X_{1}}^{4}+{k_{4}}^{2}E{X_{2}}^{4}+2k_{1}k_{3}E{X_{1}}^{3}+2k_{2}k_{4}E{X_{2}}^{3}+2k_{1}k_{2}EX_{1}X_{2}+\left( 2k_{1}k_{4}+2k_{2}k_{5} \right)EX_{1}X_{2}^{2}+\left( 2k_{1}k_{5}+2k_{2}k_{3} \right)EX_{1}^{2}X_{2}+2k_{3}k_{5}E{X_{1}}^{3}X_{2}+2k_{4}k_{5}EX_{1}X_{2}^{3}+\left( 2k_{3}k_{4}+{k_{5}}^{2} \right)EX_{1}^{2}X_{2}^{2},$$

where

$$k_{1}=A_{1},$$

$$k_{2}=\frac{TA_{2}-CA_{3}}{T-C},$$

$$k_{3}=\frac{A_{2}-A_{1}}{2C},$$

$$k_{4}=\frac{A_{3}-A_{2}}{2\left( T-C \right)},$$

$$k_{5}=\frac{A_{3}-A_{2}}{T-C}.$$

With these formulas, we can calculate $\mathrm{Var}\left( X^{*} \right)=E{X^{*}}^{2}-\left( EX^{*} \right)^{2}.$

### Appendix B, Additional simulation results

#### Comparison of Imputation Methods

We use scenario 1 with various missing rates from the main article as an example to show the difference between the imputation method 1, which only uses each subject’s own scores to impute their missing scores, imputation method 2, which uses the group average and standard deviation to impute the scores at key time points, and imputation method 3, which is the trajectory-mean imputation recommended by literature^2^. As shown in Table S1, imputation method 2 tends to have higher power than method 1, especially when the missing rate is higher. Its performance is barely influenced by increased missing rate, as the imputation at key time points using the group average is quite accurate even with more missing scores. Meanwhile, imputation method 1 tends to give bad estimates for subjects that have missing utility scores. Besides, imputation method 3 has slightly higher power than imputation method 2, though the increase is quite limited. For convenience, we use imputation method 2 by default.

**Table S1.** Power comparison of imputation methods in scenarios 1 and 2.

| $p_{\mathrm{censoring}}=p_{\mathrm{missingU}}=0.3$ | | | | | | | | | |
| --- | --- | --- | --- | --- | --- | --- | --- | --- | --- |
| $n_{1}, n_{2}$ | Imputation method 1 | | | Imputation method 2 | | | Imputation method 3 | | |
|  | $\lambda_{2}=1$ | $\lambda_{2}=0.5$ | $\lambda_{2}=2$ | $\lambda_{2}=1$ | $\lambda_{2}=0.5$ | $\lambda_{2}=2$ | $\lambda_{2}=1$ | $\lambda_{2}=0.5$ | $\lambda_{2}=2$ |
| 50 | 0.52 | 0.26 | 0.81 | 0.56 | 0.28 | 0.90 | 0.62 | 0.29 | 0.98 |
| 100 | 0.78 | 0.44 | 0.94 | 0.85 | 0.44 | 1 | 0.88 | 0.46 | 1 |
| 150 | 0.90 | 0.56 | 1.00 | 0.95 | 0.59 | 1 | 0.98 | 0.60 | 1 |
| 200 | 0.98 | 0.68 | 1.00 | 1 | 0.71 | 1 | 1 | 0.72 | 1 |
| $p_{\mathrm{censoring}}=p_{\mathrm{missingU}}=0.6$ | | | | | | | | | |
| $n_{1}, n_{2}$ | Imputation method 1 | | | Imputation method 2 | | | Imputation method 3 | | |
|  | $\lambda_{2}=1$ | $\lambda_{2}=0.5$ | $\lambda_{2}=2$ | $\lambda_{2}=1$ | $\lambda_{2}=0.5$ | $\lambda_{2}=2$ | $\lambda_{2}=1$ | $\lambda_{2}=0.5$ | $\lambda_{2}=2$ |
| 50 | 0.18 | 0.10 | 0.28 | 0.59 | 0.28 | 0.87 | 0.62 | 0.29 | 0.97 |
| 100 | 0.30 | 0.20 | 0.54 | 0.84 | 0.44 | 1 | 0.87 | 0.46 | 1 |
| 150 | 0.47 | 0.26 | 0.64 | 0.94 | 0.60 | 1 | 0.98 | 0.6 | 1 |
| 200 | 0.46 | 0.23 | 0.73 | 0.99 | 0.71 | 1 | 1 | 0.73 | 1 |

#### Exploration of the Variance Balance Factors

We use scenarios 1 and 2 from the main article as an example to show the consistency of the variance balance factors under different sample sizes. According to Table S2, the estimates of $\phi_{1}$ and $\phi_{2}$ are quite robust to the change of sample sizes, showing that the following assumed property is reasonable:

$$\mathrm{SE}\left( Q_{1} \right)=\phi_{1}\frac{\mathrm{SD}\left( {X^{*}}_{1} \right)}{\sqrt{n_{1}}},$$

$$\mathrm{SE}\left( Q_{2} \right)=\phi_{2}\frac{\mathrm{SD}\left( {X^{*}}_{2} \right)}{\sqrt{n_{2}}}.$$

**Table S2.** Average estimates based on 4000 replications.

| Scenario 1 | | | | | |
| --- | --- | --- | --- | --- | --- |
| $n_{1}, n_{2}$ | 50 | 200 | 500 | 1000 | 2000 |
| $\phi_{1}$ | 1.08 | 1.07 | 1.06 | 1.09 | 1.09 |
| $\phi_{2}$ | 1.10 | 1.12 | 1.11 | 1.10 | 1.10 |
| Scenario 2 | | | | | |
| $n_{1}, n_{2}$ | 50 | 200 | 500 | 1000 | 2000 |
| $\phi_{1}$ | 1.13 | 1.09 | 1.11 | 1.13 | 1.13 |
| $\phi_{2}$ | 1.17 | 1.15 | 1.16 | 1.14 | 1.15 |

Table S3 shows how the factor estimates change when we modify the utility missing rate in scenario 1. As the missing rate increases for the two treatment groups, both $\phi_{1}$ and $\phi_{2}$ increase. This is expected since the variance balance factors are used to bridge the difference between the variances of ${X^{*}}_{1}, {X^{*}}_{2}$ and the variances of $Q_{1}, Q_{2}$. The former is based on specified parameters only and does not involve missing data, while the latter incorporates KM estimation and utility score imputation. $\phi_{1}$ and $\phi_{2}$ can be interpreted as the scales in which extra variations are added into the HUS statistics. The more missing utility we have, the larger these scales should be.

**Table S3.** Average estimates based on 4000 replications with modified missing rates for both treatment groups in scenario 1.

| Missing rate | 0 | 20% | 40% | 60% | 80% |
| --- | --- | --- | --- | --- | --- |
| $\phi_{1}$ | 1.04 | 1.06 | 1.08 | 1.08 | 1.16 |
| $\phi_{2}$ | 1.07 | 1.10 | 1.14 | 1.16 | 1.27 |

#### Comparison of Bootstrap, Permutation and Jackknife

Table S4 shows the power of different methods with different sample sizes in scenarios 1 and 2 from the main article. Bootstrap and permutation both have similar power performance compared to the theoretical results, while bootstrap’s power tends to be slightly higher than permutation’s. Meanwhile, Jackknife’s performance is very close to bootstrap’s. We also illustrate the resampling distributions of the test statistic for one run in Figure S1. We only show the distributions of bootstrap and permutation since the distribution of Jackknife is very close to that of bootstrap. As shown in the figure, the permutation distribution is centered at 0, while the bootstrap distribution is centered at the observed test statistic, which demonstrates that permutation generates new data under the null, while bootstrap generates new data under the alternative. From the perspective of controlling type I errors in hypothesis testing, it may seem more appropriate to apply permutation. However, the bootstrap technique is widely used and can conveniently generate a confidence interval for the test statistic under the alternative.^3^ Also considering its better power performance, we recommend using bootstrap as default.

**Table S4.** Power comparison of bootstrap and permutation in scenarios 1 and 2.

| Scenario 1 | | | | |
| --- | --- | --- | --- | --- |
| $n_{1}, n_{2}$ | Theoretical | Empirical | | |
|  |  | Bootstrap | Permutation | Jackknife |
| 50 | 0.61 | 0.56 | 0.44 | 0.54 |
| 100 | 0.86 | 0.85 | 0.75 | 0.86 |
| 150 | 0.95 | 0.95 | 0.88 | 0.95 |
| 200 | 0.99 | 1 | 0.98 | 1 |
| Scenario 2 | | | | |
| $n_{1}, n_{2}$ | Theoretical | Empirical | | |
|  |  | Bootstrap | Permutation | Jackknife |
| 50 | 0.42 | 0.42 | 0.37 | 0.41 |
| 100 | 0.65 | 0.67 | 0.62 | 0.67 |
| 150 | 0.80 | 0.82 | 0.77 | 0.83 |
| 200 | 0.89 | 0.92 | 0.91 | 0.92 |

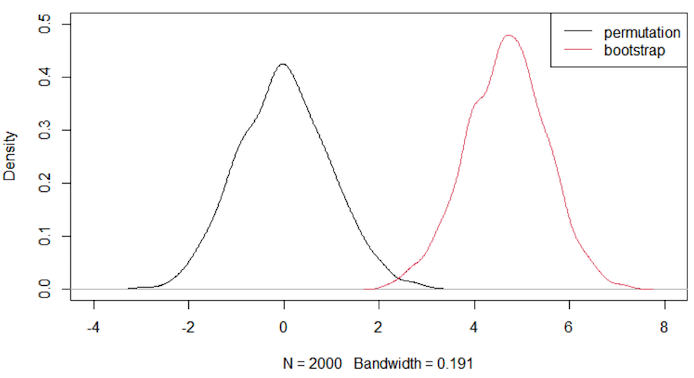

**Fig.S1.** Distributions of the test statistic based on different resampling techniques with $n_{1}=n_{2}=200$ in scenario 1.

Table S5 shows an example comparing the results of bootstrap with different numbers of iterations. The performance is very close for $B\geq500$. As a result, we recommend using $B=500$.

**Table S5.** Power comparison of bootstrap using different numbers of iterations ($B$).

| Scenario 1, bootstrap | | | | |
| --- | --- | --- | --- | --- |
| $n_{1}, n_{2}$ | $B=500$ | $B=1000$ | $B=2000$ | $B=5000$ |
| 50 | 0.44 | 0.43 | 0.43 | 0.44 |
| 100 | 0.75 | 0.75 | 0.73 | 0.76 |
| 150 | 0.88 | 0.88 | 0.87 | 0.89 |
| 200 | 0.98 | 0.99 | 0.97 | 0.97 |

#### Scenario S1 results (Two treatment Groups Only Differ in Health Utility)

For scenario S1, we assume $T=36$ and $C=12$. The base utility functions are illustrated in Figure S2. In this scenario, treatment group 1 has worse utility in the early stage of the study, but the utility recovers much faster than treatment group 2. The overall HUS of group 1 is still better than group 2. We also assume that utility scores are collected every 3 months with a 30% missing rate. As shown in Table S6, HUS is able to achieve higher power than OS-based tests, and $\lambda_{2}=2$ further increases the power. To ensure 80% power, standard HUS requires less than 200 subjects per arm.

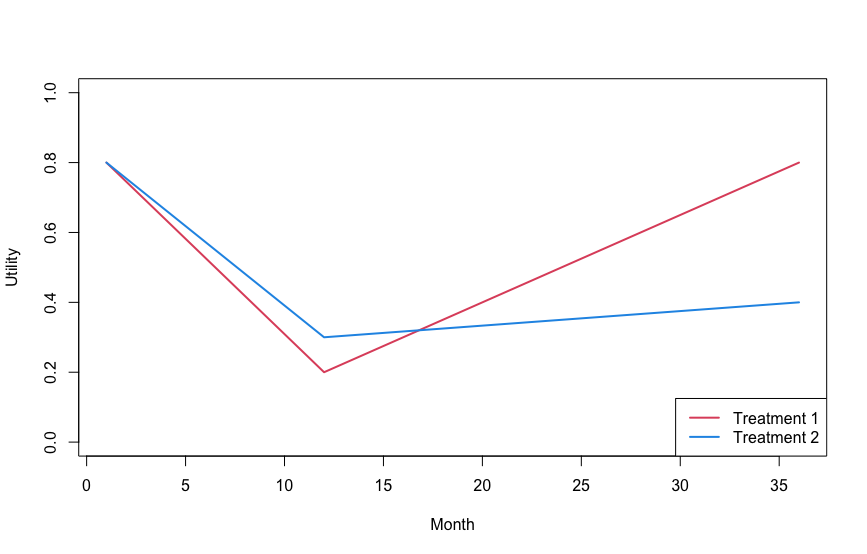

**Fig.S2.** Base utility scores for scenario S1.

**Table S6.** Power comparison for scenario S1.

| $n_{1}, n_{2}$ | HUS | | | OS | | |
| --- | --- | --- | --- | --- | --- | --- |
|  | Bootstrap | $\lambda_{2}=0.5$ | $\lambda_{2}=2$ | Sup | 5% | 10% |
| 50 | 0.38 | 0.12 | 0.84 | 0.05 | 0.04 | 0.05 |
| 100 | 0.57 | 0.22 | 1 | 0.05 | 0.05 | 0.07 |
| 150 | 0.74 | 0.32 | 1 | 0.05 | 0.06 | 0.1 |
| 200 | 0.88 | 0.32 | 1 | 0.06 | 0.04 | 0.06 |

#### Scenario S2 results (Two Groups Only Differ in Health Utility)

For scenario S2, we assume $T=36$ and each group’s base utility is flat, as illustrated in Figure S3. As shown in Table S7, our results and conclusions are similar to what we have previously. To achieve 80% power, 200 subjects per treatment group is sufficient.

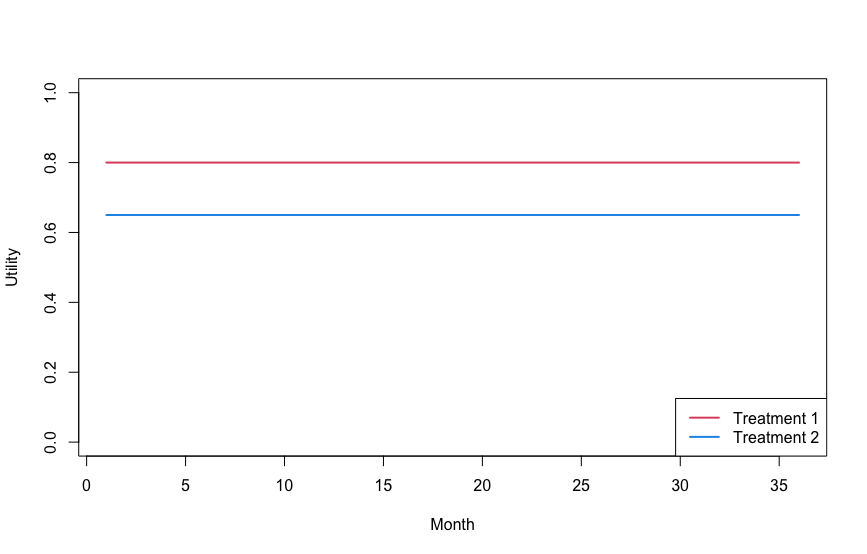

**Fig.S3.** Base utility scores for scenario S2.

**Table S7.** Power comparison for scenario S2.

| $n_{1}, n_{2}$ | HUS | | | OS | | |
| --- | --- | --- | --- | --- | --- | --- |
|  | Bootstrap | $\lambda_{2}=0.5$ | $\lambda_{2}=2$ | Sup | 5% | 10% |
| 50 | 0.38 | 0.14 | 0.76 | 0.05 | 0.04 | 0.05 |
| 100 | 0.58 | 0.26 | 0.98 | 0.05 | 0.05 | 0.07 |
| 150 | 0.74 | 0.35 | 1 | 0.05 | 0.06 | 0.1 |
| 200 | 0.88 | 0.39 | 1 | 0.06 | 0.04 | 0.06 |

#### Scenario S3 results (Two Groups Differ in Both Survival and Utility)

For scenario S3, we follow the simulation settings for scenario 1, but we set the hazard ratio as 0.8 instead of 1. We also slightly decrease the difference in health utility, as shown in Figure S4. According to Table S8, with two groups differing in OS, tests based on OS are able to obtain some power, but the power of HUS is still much higher, especially for $\lambda_{2}=1$ and $\lambda_{2}=2$. About 150 subjects per arm is sufficient for standard HUS to obtain 90% power.

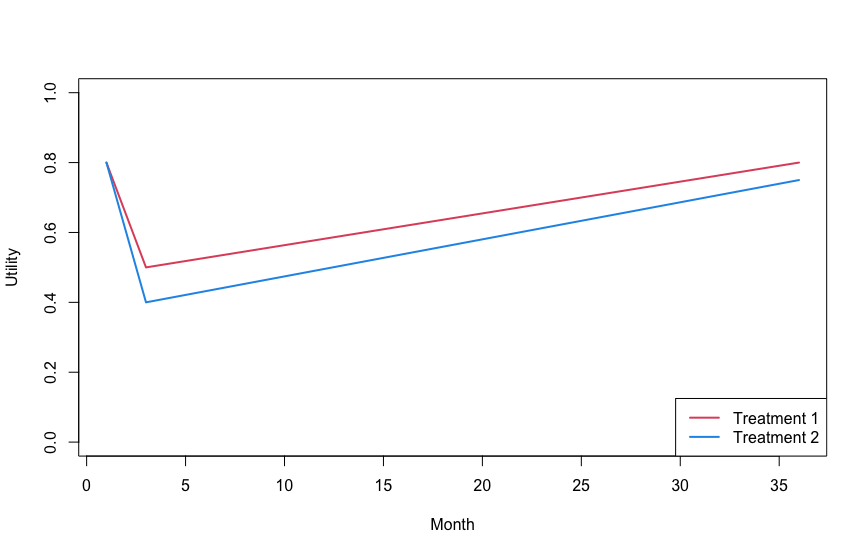

**Fig.S4.** Base utility scores for scenario S3.

**Table S8.** Power comparison for scenario S3.

| $n_{1}, n_{2}$ | HUS | | | OS | | |
| --- | --- | --- | --- | --- | --- | --- |
|  | Bootstrap | $\lambda_{2}=0.5$ | $\lambda_{2}=2$ | Sup | 5% | 10% |
| 50 | 0.48 | 0.35 | 0.74 | 0.12 | 0.14 | 0.18 |
| 100 | 0.76 | 0.56 | 0.96 | 0.22 | 0.25 | 0.34 |
| 150 | 0.91 | 0.7 | 0.98 | 0.28 | 0.38 | 0.46 |
| 200 | 0.98 | 0.84 | 1 | 0.32 | 0.42 | 0.56 |

#### Scenario S4 results (Two Groups Only Differ in Survival)

For scenario S4, we follow the simulation settings with real data estimates in the main article except that we change the base utility function of PET-CT to be the same as that of planned ND. This means the two groups only differ in OS. As shown in Table S9, when there is no difference in health utility, choosing a larger $\lambda_{2}$ decreases the power, which is why we need to be careful with $\lambda_{2}$ in practice. Meanwhile, the non-inferiority tests based on OS show higher power than HUS, which makes sense because our HUS method compares the two groups in terms of superiority rather than non-inferiority. In this scenario, tests based on HUS are similar to the superiority test based on OS, since there is only a difference in OS, and it is expected that according to the construction of rejection regions, the non-inferiority tests should have higher power than the superiority tests. As a result, OS should still be used as the primary endpoint when there is unlikely to be a difference in health utility.

**Table S9.** Power comparison for scenario S4.

| $n_{1}, n_{2}$ | HUS | | | OS | | |
| --- | --- | --- | --- | --- | --- | --- |
|  | Bootstrap | $\lambda_{2}=0.5$ | $\lambda_{2}=2$ | Sup | 5% | 10% |
| 50 | 0.12 | 0.14 | 0.12 | 0.09 | 0.1 | 0.12 |
| 100 | 0.17 | 0.18 | 0.18 | 0.14 | 0.17 | 0.22 |
| 282 | 0.3 | 0.32 | 0.3 | 0.27 | 0.36 | 0.44 |
| 500 | 0.41 | 0.44 | 0.37 | 0.36 | 0.5 | 0.64 |
